# B.1.617.2 SARS-CoV-2 (Delta) variant is associated with increased risk of hospitalization and death compared with B.1.1.7 SARS-CoV-2 (Alpha) variant

**DOI:** 10.1101/2022.01.21.22268602

**Authors:** Eduardo Freire Rodrigues, Joana Moreno, Pedro Pinto Leite, Pedro Casaca, Baltazar Nunes, João Paulo Gomes, Rita Ferreira, Joana Isidro, Vítor Borges, Luís Vieira, Sílvia Duarte, Carlos Sousa, José Pedro Almeida, Luís Menezes, Dora Vaz, Andreia Leite, André Peralta-Santos

## Abstract

**Introduction:** The B.1.617.2 variant (Delta) was associated with increased transmissibility and lower vaccine effectiveness than the B.1.1.7 variant (Alpha). However, the effect of the B.1.617.2 variant on disease severity remains unclear. This study aims to assess whether infection with the B.1.617.2 variant was associated with a higher risk of serious illness, compared with other co-circulating variants, measured through hospitalization and death by COVID-19 in Portugal.

**Methods:** We conducted a matched cohort study in adult individuals diagnosed with SARS-CoV-2/COVID-19 infection between March 29 and August 1, 2021. Cases were individuals with a positive PCR test notified to the surveillance system. SARS-CoV-2 variants were classified first by genomic sequencing (WGS) and, if this information was unavailable, by detecting the S gene target failure.

Delta (B.1.617.2) and Alpha (B.1.1.7) cases were matched on the week of diagnosis at a 1 to k ratio (k being the maximum number of unexposed available in that week) to maximize the inclusion of unexposed, using the nearest-neighbor algorithm. The hazard risk and 95% confidence intervals of hospitalization and death among those infected with the Delta (B.1.617.2) variant vs. Alpha (B.1.1.7) was estimated using a Cox proportional hazards model, adjusting for confounding for sex, age, and vaccination status.

**Results:** A total of 2,778 cases were included in the study. Of the total, 1 742 (68%) were identified as B.1.617.2 variant cases and 3 629 (32%) as B.1.1.7 variant. Within the B.1.1.7 variant cases 106 (2.9%) were hospitalized, and 110 (6.3%) within the B.1.617.2 variant cases. A total of 29 deaths were reported, 8 (0.2%) in patients infected with B.1.1.7 variant and 21 (1.2%) in patients with the B.1.617.2 variant. The confounding adjusted risk of hospitalization, in persons infected with the B.1.617.2 variant was 2.44 (95%CI 1.85; 3.20) times higher than the risk of hospitalization among B.1.1.7 variant cases, and the confounding-adjusted risk of death for B.1.617.2 variant cases was 5.20 (95%CI 2.20; 12.29) times higher than the risk of death in patients infected by B.1.1.7 variant.

**Conclusion:** The B.1.617.2 variant is associated with an increased risk of hospitalization and death compared with the B.1.1.7 variant.

## Background

The adaptation processes of SARS-CoV-2, characterized by different mutations, are a challenge to contain the pandemic impact on health systems and population health. Most mutations have little influence on the virus’ ability to cause disease. However, the estimated mutation rate of SARS-CoV-2 is more than four times higher than the seasonal Influenza HA1 subunit (1). Depending on where these mutations are in the genetic chain, they may change the virus’ properties. When mutations yield a selective advantage by increasing the transmissibility or the ability to evade the host immune response or increasing disease severity, they are classified as Variants of Concern (VOC) (2).

The B.1.1.7 SARS-CoV-2 (also known as Alpha variant) is characterized by several spike protein changes being the deletion 69-70 the one responsible for changes in S-gene detection on RT-PCR assays (3, 4). Since its first detection, the spread of this variant to the UE/EEA countries was fast, becoming the most prevalent variant in many countries as Portugal, in the first months of 2021 (4).

The dominant variant in most UE/EEA countries, until December 2021, was the lineage B.1.617.2 of SARS-CoV-2, also known as the Delta variant. This variant was identified in India in December 2020, was classified as VOC in May 2021. It is associated with increased transmissibility and reduced antibody neutralization (5, 6). In Portugal, the B.1.617.2 variant was first identified at the end of April 2021 and became dominant in July, representing 90% of all sequenced cases. Unlike variant B.1.1.7, the B.1.617.2 variant is not characterized by the deletion 69-70 in the spike protein.

Vaccines are currently the best solution to control the impact of the SARS-CoV-2 pandemic and reduce the burden on healthcare institutions’ capacity and mortality (6). Nevertheless, the emergence of new SARS-CoV-2 variants with increased immune evasion may reduce the vaccine effectiveness (7) and cause stress on health systems. The Omicron VOC emergence in November 2021 (8) is a testament to the vaccine effectiveness (9, 10).

Vaccine effectiveness studies have shown that current vaccines effectively prevent symptomatic infection and disease that leads to hospitalization and reduce case-fatality rate (11). Recent evidence has shown a decrease in vaccine effectiveness against the B.1.617.2 variant compared to the B.1.1.7 variant, both for infection prevention, disease severity and death (5, 12, 13).

Disease severity (rate of hospitalizations and deaths) of VOC is critical to understand the potential strain of the pandemic on health systems. Few studies have addressed the associated risk of hospitalization among patients infected with the B.1.617.2 variant compared to patients infected with the B.1.1.7 variant. Two studies conducted in England (14) and Scotland(15), which directly compared patients with the B.1.617.2 variant with B.1.1.7 patients, estimated that the risk of hospitalization admission was 2.26 and 1.85 higher in B.1.617.2 variant patients, respectively. Another study performed in the United States compared cases from B.1.617.2 variant with other non-VOC or variant of interest lineages also found a higher risk of hospitalization in patients infected with B.1.617.2 variant (Hazard Ratio 2.35). Nevertheless, a study in Norway did not find a higher risk of hospitalization associated with the B.1.617.2 variant (adjusted relative risk of 0.97 95%CI 0.76–1.23) (16).

As existing studies are not conclusive regarding the role of the B.1.617.2 variant in the risk of hospitalization and death, we decided to carry out this study to provide more evidence on this issue. This study intends to evaluate if B.1.617.2 variant is associated with a higher risk of severe disease, hospitalization and death, and the effect modification of vaccination status on severe disease in a context where this variant is dominant.

## Methods

### Study design

We designed a matched cohort study to assess the risk of hospitalization and the risk of death of the individuals infected with Delta (B.1.617.2) variant compared to those infected with Alpha (B.1.1.7) variant. The study population was adult individuals (>18 years old) diagnosed with SARS-CoV-2 infection/COVID-19 by nasopharyngeal swab tested with RT-PCR from March 29, 2021 (ISO week 13) to August 1, 2021 (ISO week 30) notified through the laboratory service of surveillance system (SINAVE). We included individuals with samples classified either by whole-genome sequencing or Spike Gene Target Failure (SGTF). Both symptomatic and asymptomatic individuals were included. The individuals were matched on the week of diagnosis.

### Sample collection and variant classification

We used two different methods to identify which samples were B.1.1.7 or B.1.617.2 variants. First, we used information from a nationwide network group of laboratories (UNILABS) that performs RT-PCR tests for SARS-CoV-2 using Thermofisher TaqPath assay, targeting three regions of the SARS-CoV-2 genome: ORF1ab, N and S genes. Their samples are classified as B.1.1.7 or non-B.1.1.7, according to the amplification of S-gene. Secondly, we use Whole-Genome Sequencing (WGS) data provided by the National Health Institute (INSA) that routinely performs sequencing on random samples notified to SINAVE. In case of misclassification (e.g., different classification from WGS and SGTF), we accepted the WGS result as final due to higher accuracy(17).

### Outcomes: hospitalization and death

We defined hospitalization as any admission to a public hospital with the primary diagnosis being COVID-19 (ICD-10 *code U*.*071*), obtained through the Central Hospital Morbidity Database, in the following 15 days after the confirmed diagnosis of SARS-CoV-2 infection/COVID-19. The Central Hospital Morbidity Database gathers data from all public hospitals in Portugal, which account for most hospitals in Portugal and the overwhelming majority of hospitals admitting inpatients with COVID-19.

A COVID-19 death was defined as any record of death on the national Death Certificate Information System (SICO) with COVID-19 as the primary cause of death (ICD-10 *code U*.*071*) according to the WHO classification (18). SICO platform allows the issuance of a death certificate for each person who dies in Portugal.

### Vaccination status and demographic covariates

We obtained the COVID-19 vaccination status through the electronic national vaccination register (VACINAS). Vaccination exposure was indexed at the date of COVID-19 diagnosis and classified as: (i) unvaccinated: no register of vaccine administration prior to diagnosis; (ii) partial vaccination: SARS-CoV-2 infection/COVID-19 diagnosis anytime between the uptake of the first dose of a COVID-19 vaccine (ChAdOx1, BNT162b2 or mRNA-1273) and up to 14 days after second dose uptake; and (iv) complete vaccination: 14 or more days following second dose vaccine uptake. Information about age, sex and date at diagnosis was routinely collected on surveillance system SINAVE and was extracted from it. Age categories were created accordingly to the functional form of the variable based on the Martingale residuals.

### Data source and record-linkage

We used data from different databases: SINAVE is the national epidemiological and laboratory surveillance platform for SARS-CoV-2 infections, SICO the electronic death certificate platform, VACINAS the electronic vaccination platform, BDMH the hospital admission platform of the National Health System, data from sequencing was provided by the National Institute of Health. Data were extracted on September 17, 2021. Record-linkage was performed using a unique identifier (national health service number) to exclude duplicates. Duplicates or suspected reinfection were excluded.

### Matching

Matching was performed based on the week of SARS-CoV-2 infection diagnosis using variable ratio matching, at a ratio of 1 to k, where k is the maximum number of B.1.1.7 cases available on the respective week. Additionally, to optimize the use of controls in periods of B.1.617.2 dominance that resulted in a scarcity of controls for matching, we used a nearest-neighbor algorithm provided with the “Matchit”(19) CRAN package.

### Statistical Analysis

We compared the characteristics of SARS-CoV-2 cases using central tendency and dispersion measures for continuous variables and absolute and relative frequencies for categorical variables. The B.1.617.2 cases were considered exposed, while B.1.1.7 cases were considered the reference group.

The confounding-adjusted hazard ratio of hospitalization and death, with 95% confidence intervals (CI), was calculated using a proportional hazards Cox model, adjusted for sex, age, and vaccination status (equation 1).

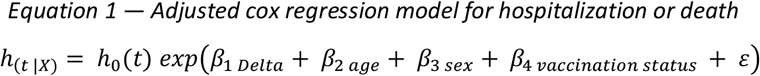

Where *h*_(*t* |*X*)_ is the hazard at the time t of the outcome hospitalization, and of death, *h*_0_(*t*) is the baseline hazard for the outcome, exp (*β*_1 *Delta*_) the hazard ratio for the outcome being infected with delta variant compared with alpha, exp (*β*_2 *age*_) are the hazard ratios for the outcome being 51 to 50 years old or 50 or older compared with being 18 to 40 years old, exp (*β*_3 *sex*_) the hazard ratio for the outcome being a female compared with being male, exp (*β*_3 *vaccination status*_) are the hazard ratio for the outcome being partially vaccinated or fully vaccinated compared with unvaccinated with, *ε*is the error term.

To check if the vaccination status was an effect modifier of the association between variant and severity, we added an interaction term between the variant and vaccination status to the model. We stratified the analysis by unvaccinated and vaccinated (partial and complete). We tested the proportionality of risk assumptions using visual inspection of scaled Schoenfeld residuals (20). We used the survival package to do the proportional hazards Cox model (21).

Due to the nature of the outcomes and their national availability, our analysis assumed a type I right censoring, thus using a fixed end of follow-up date defined as August 31, 2021. Analyses were based on a complete case analysis. All tests were two-sided, and a P-value < .05 was considered statistically significant. All statistical analyses were performed on R® using R Studio version 1.4 (22).

### Ethical Statement

The genomic surveillance of SARS-CoV-2 in Portugal is regulated by the Assistant Secretary of State and Health Executive Order (Despacho n.º 331/2021 of January 11, 2021). The research on genomic epidemiology of SARS-COV-2 received the clearance of the Ethics Committee of INSA on March 30, 2021.

## Results

### Enrolment and matching

From March 29 through August 1, 2021, we identified through WGS or TGSF with successful record linkage 18 396 SARS-CoV-2 cases in Portugal. A total of 5 371 participants were included in the study. 1 742 (68%) were identified with B.1.617.2 variant and matched with 3 629 cases (32%) identified with B.1.1.7 variant.

Most of the study participants were women (51.7%), the mean age was 41.9 (SD=15.8) years (supplementary material S1), and unvaccinated (82.3%). The mean length of stay in the hospital was 10 (SD=8.8) days. The characteristics of the patients at baseline were balanced between the groups (Table 1), mainly the age, except for the vaccination status where the patients infected with B.1.617.2 variant showed a higher vaccination coverage both for any vaccination status (23.8% versus 14.8%), but also for partial and complete vaccination coverages.

**Table 1.** Sociodemographic and clinical characteristics of the study sample, according to the identified SARS-CoV-2 variant after matching by week of diagnosis

|  | **B.1.1.7**<br>**Alpha**<br>**n (%)** | **B.1.617.2**<br>**Delta**<br>**n (%)** |
| --- | --- | --- |
| **Total** | 3 629 | 1 742 |
| **Sex** |  |  |
| Male | 1 713 (47.2) | 880 (50.5) |
| Female | 1 916 (52.8) | 862 (49.5) |
| **Age** |  |  |
| 18-40 | 1 734 (47.8) | 907 (52.1) |
| 41-50 | 801 (22.1) | 269 (21.2) |
| 51+ | 1 094 (30.1) | 466 (26.8) |
| **Vaccination Status** |  |  |
| Unvaccinated | 3 092 (85.2) | 1 327 (76.2) |
| Partial | 440 (12.1) | 278 (16.0) |
| Complete | 97 (2.7) | 137 (7.9) |
| **Hospitalization** |  |  |
| No | 3 523 (97.1) | 1 632 (93.7) |
| Yes | 106 (2.9) | 110 (6.3) |
| **Length of Stay**<br>**(hospitalization)**<br>**Mean (SD)** | 10.3 (8.6) | 10.0 (9.1) |
| **Death** |  |  |
| No | 3 621 (99.8) | 1 721 (98.8) |
| Yes | 8 (0.2) | 21 (1.2) |

### Outcomes

We observed 216 hospitalizations and 29 deaths between March 29 and August 1, 2021. There were 226 hospitalizations, 106 (2.9%) among patients infected with the B.1.1.7 and 110 (6.3%) with the B.1.617.2 variant. A total of 29 deaths were reported, 8 (0.2%) in patients infected with B.1.1.7 variant and 21 (1.2%) in patients with the B.1.617.2 variant.

For the outcome hospitalization, the mean duration of follow-up was 51.2 (SD=23.3) days for B.1.617.2 and 71.8 (SD=28.4) days for B.1.1.7. For the outcome death, the mean duration of follow-up was 53.9 (SD=20.8) days for B.1.617.2 and 73.9 (SD=26.4) days for B.1.1.7. We observed longer follow-ups on patients infected with the B.1.1.7 variant due to its dominance at the start of the study period, followed by a transition to dominance by the B.1.617.2 variant.

### Hospitalization

The crude cumulative hospitalization probability at the 15^th^ day after a diagnosis of SARS-CoV-2 infection was 2.8% (95%CI 2.3% to 3.3%) for patients infected with the B.1.1.7 variant and 6.2% (95%CI 5.1 to 7.3) for patients infected with the B.1.617.2 variant. There were no hospitalization events observed following the 39^th^ day after a diagnosis of COVID-19. Regarding hospital admissions, the unadjusted hazard ratio of hospitalization was 2.18 (95%CI 1.67 to 2.85). The adjusted age, sex, vaccination status and date of diagnosis for HR for hospitalization for the B.1.617.2 variant was 2.44 (95%CI 1.85 to 3.20) times the risk of B.1.1.7 variant (table 2). When we tested for the interaction between the variant and the vaccination status, the HR coefficients were statistically significant for the hospitalization outcome. Stratified by vaccination status, the increased risk for hospitalization for Delta patients is higher in the unvaccinated strata, HR 2.78 (95%CI 2.02 to 3.82) than for vaccinated participants HR 1.59 (95%CI 0.94 to 2.69), however we cannot exclude that the risk of hospitalization could be same, between patients infected with Alpha and Delta, in vaccinated participants, see table 3.

**Table 2.** Hospitalization and death risk of B.1.617.2 (Delta) versus B.1.1.7 (Alpha)

| Primary Outcomes | B.1.1.7, Alpha (n=3 629) |  | B.1.617.2, Delta (n=1 742) |  | B.1.617.2, Delta versus B.1.1.7, Alpha |  |  |  |
| --- | --- | --- | --- | --- | --- | --- | --- | --- |
|  | No. Events | Incidence Rate (per 1000 cases) | No. Events | Incidence Rate (per 1000 cases) | Crude HR (95% CI) | p-value | Adjusted <sup>a</sup> HR (95% CI) | p-value |
| **Hospitalization** | 106 | 29.20 | 110 | 63.14 | 2.18 (1.67 to 2.85) | <0.001 | 2.44 (1.85 to 3.20) | <0.001 |
| **Death** | 8 | 2.20 | 21 | 12.05 | 5.01 (2.19 to 11.44) | <0.001 | 5.20 (2.20 to 12.29) | <0.001 |

a.adjusted for age, sex, vaccination status; HR: hazard ratio

**Table 3.** Hospitalization and death risk of B.1.617.2 (Delta) versus B.1.1.7 (Alpha) stratified by vaccination status

|  | B.1.617.2, Delta versus B.1.1.7, Alpha<br>Unvaccinated |  | B.1.617.2, Delta versus B.1.1.7, Alpha<br>Vaccinated (Partial and Complete) |  |
| --- | --- | --- | --- | --- |
| Primary Outcomes | Adjusted <sup>a</sup><br>HR (95% CI) | *p-value* | Adjusted <sup>a</sup><br>HR (95% CI) | *p-value* |
| Hospitalization | 2.78 (2.02 to 3.82) | <0.001 | 1.59 (0.94 to 2.69) | 0.08 |
| Death | 8.25 (2.53; 26.93) | <0.001 | 3.39 (1.06; 10.84) | <0.05 |

a.adjusted for age, sex, vaccination status; HR: hazard ratio

### COVID-19 Death

The crude cumulative mortality probability at the 30^th^ day after a diagnosis of SARS-CoV-2 after the COVID-19 diagnosis the cumulative mortality probability was 0.2% (95%CI 0.1% to 0.4%) for B.1.1.7 and 1.1% (95%CI 0.6% to 1.6%) for B.1.617.2 infections. There were no death events observed after the 47^th^ day following COVID-19 diagnosis. As to mortality, the unadjusted hazard ratio of death was 5.01 (95%CI 2.19 to 11.44). The adjusted HR was 5.20 (95%CI 2.20 to 12.29) for the B.1.617.2 variant compared with the B.1.1.7 variant (see table 2). The HR coefficients for the adjusted models are available in the supplementary material.

When we tested for the interaction between the variant and the vaccination status, the HR coefficients were not statistically significant for the death outcome, however, we stratified to compare estimates. Stratified by vaccination status, the increased risk for hospitalization for Delta patients is higher in the unvaccinated strata, 8.25 (95%CI 2.53 to 26.93) than for vaccinated participants 3.39 (95%CI 1.06 to 10.84) with higher uncertainty, see table 3.

We found no significant violation of the proportionality of hazards assumptions (supplement materials for more information).

## Discussion

We report twice the risk of COVID-19 hospitalization in patients infected with Delta (B.1.617.2) variant when compared to the Alpha (B.1.1.7) variant patients. Moreover, the risk of death in the Delta (B.1.617.2) variant patients was five times the risk of death in Alpha (B.1.1.7) variant patients.

Previous studies have reported similar results for the risk of hospitalization using diverse study designs. We report very similar estimates of the hazard ratio of hospitalization, adjusted HR of 2.44 (95%CI 1.85 to 3.20) compared to the England cohort study (14) HR 2.26 (IC95% 1.32 to 3.89) using samples with lineage identification through WGS. Furthermore, our results are also in line with the findings of the Scotland study (15) that assessed hospitalization risk with samples with lineage identification through TGSF, but a higher magnitude was found in our study HR 1.85 (95%CI: 1.39 to 2.47). The similitude between the relative risk of hospitalization estimated between our study and the England cohort study strengthens the evidence of the increased risk by the Delta variant when compared with Alpha variant cases.

To the best of our knowledge, we provide for the first time estimates of HR for mortality comparing Delta B.1.617.2 variant versus Alpha B.1.1.7 variant in a cohort study. We estimate a hazard risk of death 5.20 (95%CI 2.20 to 12.29) for the B.1.617.2 variant compared with B.1.1.7 variant cases. These estimates are based on a small number of events and hence have low precision. However, these results are supported by cellular studies(5, 23, 24) that show a higher affinity of Delta B.1.617.2 variant for lower respiratory tract cells and thus a higher probability of severe disease and death.

We report a statistically significant interaction between variant and vaccination status for the hospitalization outcome. Stratified by vaccination status we found that the risk for hospitalization and death is consistently higher for Delta compared with Alpha, however the risk has a higher magnitude for unvaccinated participants. Moreover, for vaccinated participants the precision was too low to determine whether the risks of hospitalization were higher or similar for participants with the delta variant compared with participants with the alpha variant. This is consistent with other studies that reported similar findings (14). The risk of death is consistently higher for Delta in unvaccinated and vaccinated participants, and the interaction between variant and vaccination status was not significant.

We designed a retrospective cohort matched by the week of diagnosis design to minimize the effect of time and time-associated cofounders on risk estimates such as, e.g., bed occupancy, availability of hospitals for further admissions or vaccination coverage. Even with a variable delta to alpha ratio approach for comparator selection, this design reduced the sample size. That reduced our power, particularly for rare events, as in the mortality analysis and stratified analysis by vaccination status. However, this approach minimized biases and provided more robustness to our estimates by mitigating the effect of time-dependent confounders.

Our study benefits extensively from the secondary use of administrative and health-related data sources. These systems (SARS-CoV-2 cases, hospitalization, death, vaccination registers) and unique identifiers allow researchers to answer complex investigation questions in shorter periods and fewer resources. However, secondary use of administrative data encompasses several limitations. First and foremost, hospital admission data can have delays due to coding processes, which would significantly affect participants diagnosed closer to the end of the study period. As the B.1.617.2 variant has become dominant halfway from the end study, it can be argued that this delay would lead to an underestimation of the number of events in this group and by consequence an underestimation of the HR of death and hospitalization. We have mitigated this factor by extracting data on hospital admissions almost two months after the end of the study period.

The selection of samples for WGS and SGTF was independent of the VOC type and severity, e.g., hospitalization or death, making the risk of differential selection bias residual. Our study sample showed a skewness towards younger ages, and this may be explained by the fact that our cases were collected mainly through ambulatory laboratories. At the same time, older (80+) individuals are expected to be more frequently diagnosed by hospital laboratories. While the cumulative risk of hospitalization and death could be underestimated, it is unlikely that the age distribution in the study would bias the variant HR estimates.

We cannot exclude residual confounding in the study, our study did not account comorbidities. Patients infected with Delta variant for the same age could be sicker (immunocompromised, higher burden of comorbidities) and that affected the risk of hospitalization and death disproportionately for Delta patients. Other variables, such as such as place of residence are unlikely to be confounders. On the one hand the probability of differences by variant is low, as the B.1.617.2 variant became dominant similarly throughout the country. On the other hand differences in hospital admission by region due to access conditions is not expected as Portugal has a mainly national healthcare system with good regional coverage and regional availability (25).

Additionally, there was no information available on the presence and type of symptoms at the time of diagnosis. As such, we cannot describe the proportion of symptomatic and asymptomatic infections between patients infected with B.1.1.7 and with B.1.617.2. However, the risk of hospitalization and death is not influenced by the distribution and quality of symptoms and is a mediator effect and not a confounder. All the patients with symptoms and meeting the criteria for hospitalization in national guidelines should be hospitalized.

Misclassification of variant is a potential limitation because we used S gene no amplification to identify the variant. This method has been used before, mainly in the Scottish study (15), and misclassification is minimized because we used sequencing data whenever present.

## Conclusion

This study supports the hypothesis of higher severity of Delta variant. We report a risk of COVID-19 hospitalization that was double (HR 2.44, 95%CI 1.85 to 3.20) for participants infected with the Delta variant when compared to the Alpha variant and risk of death in the Delta variant participants is five times (HR 5.20 95%CI 2.20 to 12.29) than for Alpha participants.

For unvaccinated patients, the HR risk for hospitalization and death is consistently higher for Delta than Alpha. However, we cannot exclude that the risk of hospitalization in vaccinated patients could be identical between patients infected with Alpha and Delta, HR 1.59 (95%CI 0.94 to 2.69). The risk of death is still higher for Delta than Alpha, in vaccinated patients HR 3.39 (95%CI 1.06 to 10.84).

## Data Availability

All data produced in the present study are available upon reasonable request to the authors

