## Supplementary material for "B.1.617.2 SARS-CoV-2 (Delta) variant is associated with increased risk of hospitalization and death compared with B.1.1.7 SARS-CoV-2 (Alpha) variant"

**Supplementary Materials**

**Figure S1 — Age density per virus variant after matching**

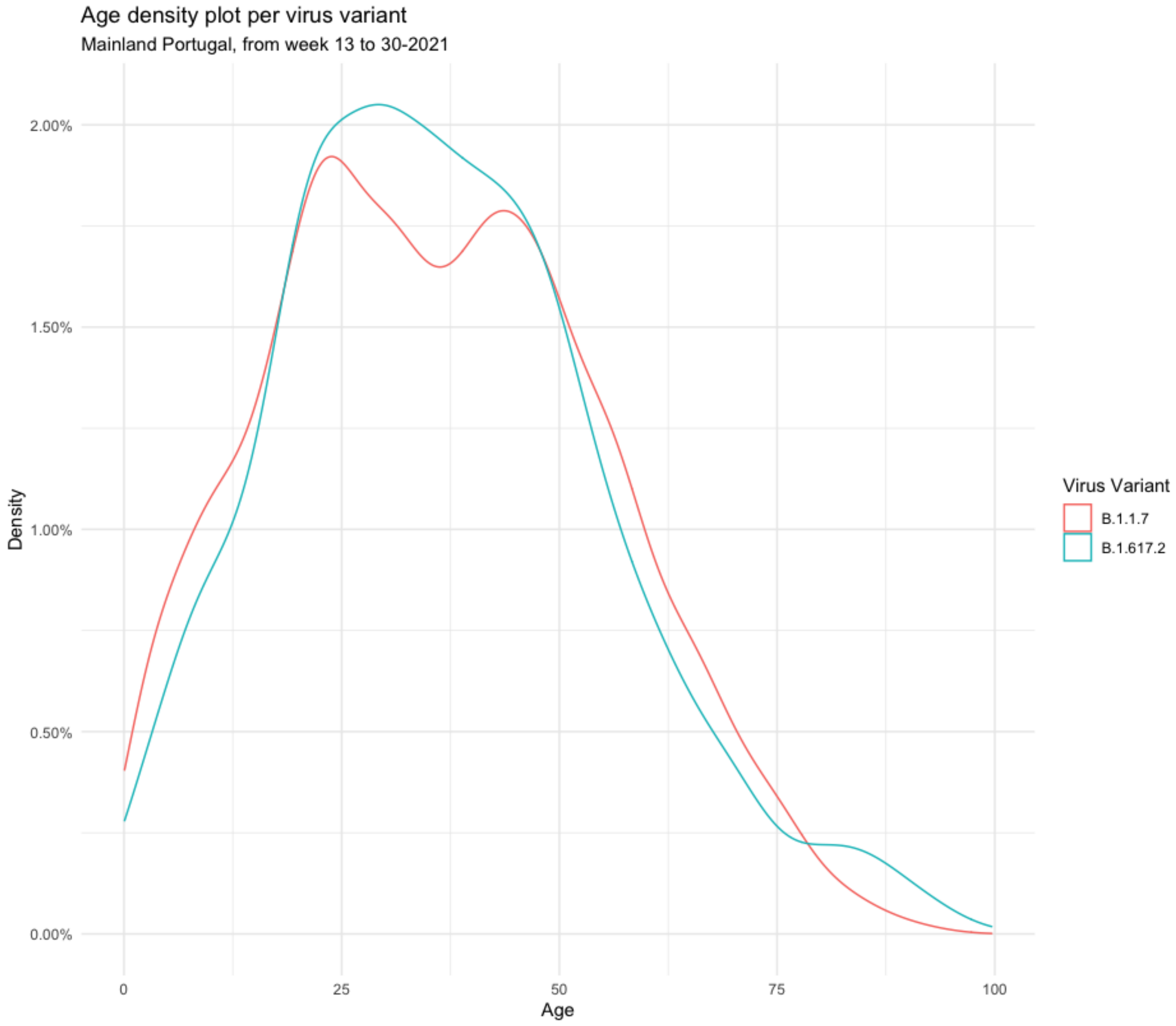

*Table S 1. Hazard Ratio for hospitalization in the adjusted model*

| Variable | HR hospitalization | P-value | Low CI | High CI |
|---|---|---|---|---|
| Alpha (B.1.1.7) | Ref. | | | |
| Delta (B.1.617.2) | 2.44 | 0.000 | 1.85 | 3.21 |
| Age [18-40) | Ref. | | | |
| Age (40,50] | 1.99 | 0.001 | 1.31 | 3.02 |
| Age (50,120] | 5.30 | 0.000 | 3.72 | 7.56 |
| Sex Female | Ref. | | | |
| Sex Male | 1.56 | 0.001 | 1.19 | 2.05 |
| Vaccination status \| Unvaccinated | Ref. | | | |
| Vaccination status \| Partial | 0.78 | 0.185 | 0.55 | 1.12 |
| Vaccination status \| Full | 0.68 | 0.200 | 0.38 | 1.23 |

*Table S 2. Hazard Ratio for death in the adjusted model*

| Variable | HR Death | P-value | Low CI | High CI |
|---|---|---|---|---|
| Alpha (B.1.1.7) | Ref. | | | |
| Delta (B.1.617.2) | 5.20 | 0.000 | 2.20 | 12.29 |
| Age [18-40) | Ref. | | | |
| Age (40,50] | 2.35 | 0.546 | 0.15 | 37.62 |
| Age (50,120] | 44.10 | 0.000 | 5.75 | 338.23 |
| Sex Female | Ref. | | | |
| Sex Male | 1.12 | 0.776 | 0.52 | 2.38 |
| Vaccination status \| Unvaccinated | Ref. | | | |
| Vaccination status \| Partial | 0.88 | 0.791 | 0.35 | 2.20 |
| Vaccination status \| Full | 1.80 | 0.261 | 0.65 | 5.01 |

**Figure S2 — Covariate proportionality**

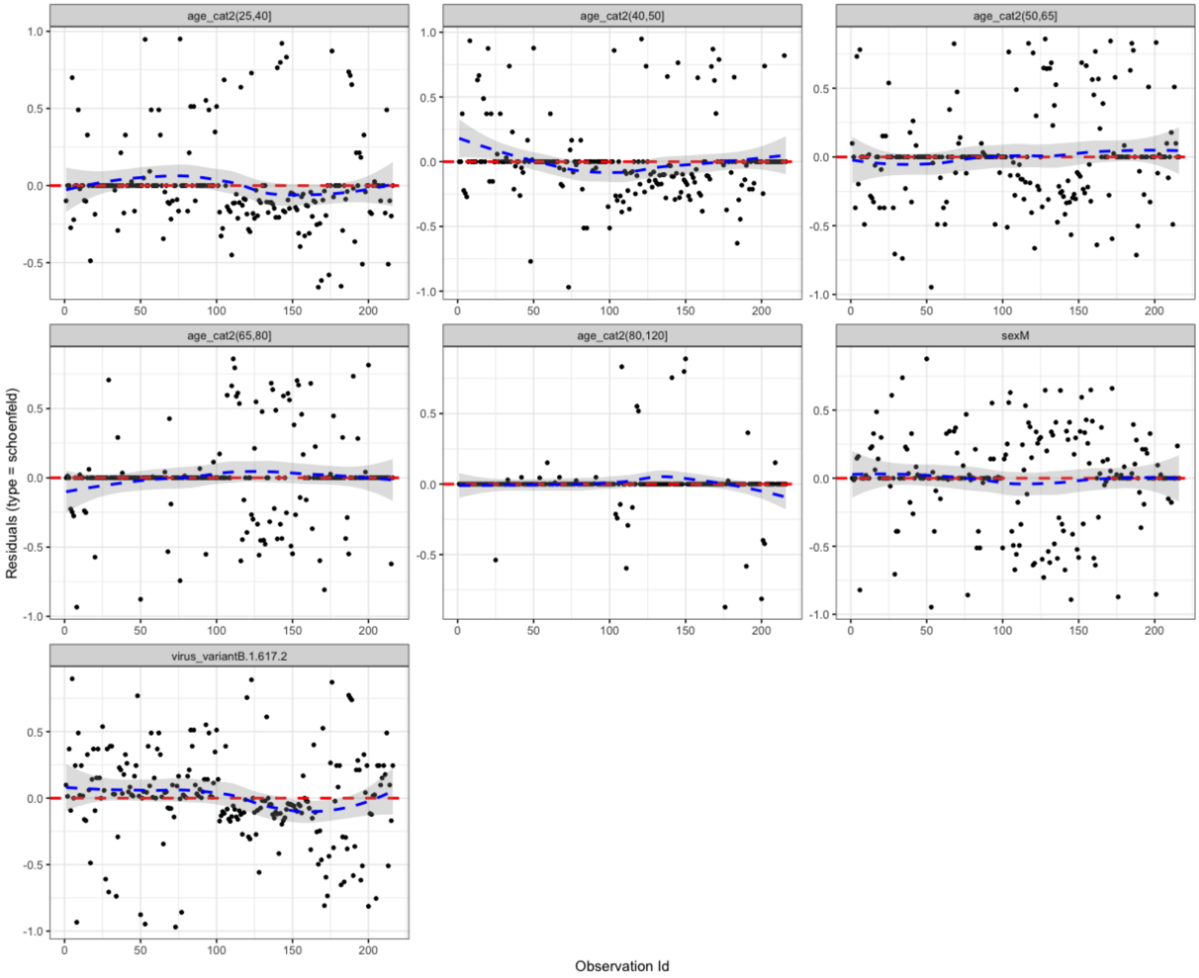
